# Death associated with coronavirus (COVID-19) infection in individuals with severe mental disorders in Sweden during the early months of the outbreak – a exploratory cross-sectional analysis of a population-based register study

**DOI:** 10.1101/2020.09.14.20193987

**Authors:** Martin Maripuu, Marie Bendix, Louise Öhlund, Micael Widerström, Ursula Werneke

## Abstract

**Background:** Individuals with severe mental disorder (SMD) have a higher risk of somatic comorbidity and mortality than the rest of the population. We set up a population-based study to assess whether individuals with SMD had a higher risk of death associated with a COVID-19 infection (COVID-19 associated death) than individuals without SMD.

**Methods:** Exploratory analysis with a cross-sectional design in the framework of a population-based register study covering the entire Swedish population. The Swedish Board for Health and Welfare (Socialstyrelsen) provided anonymised tabulated summary data for further analysis. We compared numbers of COVID-19 associated death in individuals with SMD (cases) and without SMD (controls). We calculated the odds ratio (OR) for the whole sample and by age group and four potential risk factors, namely diabetes, cardiovascular disease, hypertension, chronic lung disease.

**Results:** The sample comprised of 7,923,859 individuals, 103,999 with SMD and 7,819,860 controls. There were 130 (0.1%) COVID-19 associated deaths in the SMD group and 4945 (0.06%) in the control group, corresponding to an OR of 1.98 (CI 1.66-2.35; p < 0.001). The odds were fourfold in the age group between 60 and 79 years. Cardiovascular diseases increased the odds by 50%. Individuals with SMD without any of the risk factors under study had three-folds odds of COVID-19 associated death.

**Conclusion:** Our preliminary results suggest that individuals with SMD are a further group at increased risk of COVID-19 associated death. The factors contributing to this increased mortality risk require clarification.

## Introduction

Severe mental disorders (SMD), such as bipolar or psychotic disorders are associated with premature mortality. SMD has a substantial impact on life expectancy, which may be shortened by 10 to 20 years.^1-3^ Somatic disorders account for at least 50% of this shortened life expectancy.^3^ Cardiovascular conditions are the main cause of premature death.^3-5^ Infectious diseases also seem to contribute to a shortened life expectancy in individuals with SMD.^1,2^ It currently remains unclear whether individuals with severe SMD have an increased risk of death associated with coronavirus SARS-CoV-2 (COVID-19) infection. The question is important, since individuals with SMD may have an excess risk of factors linked to an adverse outcome of COVID-19 infection. Such risk factors include cardiovascular conditions, diabetes, chronic respiratory disease, hypertension and obesity.^6,7^ At the same time, individuals with SMD have less access to somatic health care.^5,8^ Therefore, there are reasons to believe that individuals with SMDs experience a higher mortality associated with COVID-19 infection than the rest of the population. If this assumption held true, individuals with SMD would join the category of individuals at increased risk of an adverse clinical course of COVID-19 infection, along with people who are older, obese or who have pre-existing somatic conditions. We set up a national register study to examine this hypothesis. In order to be able to make health professionals aware of this potential risk group as soon as possible, we conducted a first exploratory analysis, which we report here.

### Aim

To assess the risk of death associated with COVID-19 infection (COVID-19 associated death) in individuals with SMD. We tested the following hypothesis: Individuals with SMD have a higher risk of COVID-19 associated death than individuals without SMD (reference population).

## Methods

### Study design

The current study is part of a longitudinal population-based register study based on the Swedish National Patient Register, the Swedish National Death Register, and the Swedish Prescribed Drug Register. All registers are held by the Swedish Board for Health and Welfare (Socialstyrelsen). For this exploratory analysis, we chose a cross-sectional design, linking the Swedish National Patient Register and the Swedish National Death Register. The data manager at the Swedish Board for Health and Welfare linked the respective registers through the unique personal identification number (Swedish personal number). Anonymised tabulated summary data was made available to the research team for further analysis. We checked our whole method against the Strobe checklist (Appendix 1).^9^

### Data Sources

The Swedish National Patient Register is based on diagnoses for both in and outpatient care in specialised medicine (secondary care). Diagnoses from general practitioners (primary care) are not included in this register. The Swedish National Death Register includes all Swedish persons that have died. The cause of death is established in either primary or secondary care, depending on where the death has occurred. The Swedish Prescribed Drug Register includes all prescribed and purchased drugs in Sweden, irrespective of level of care.

### Sample

We included the whole Swedish population of at least 20 years of age by 31 Dec 2019. We defined individuals with a diagnosis of SMD as cases and all other individuals as controls. As the sample covered the whole adult Swedish population, all individuals fell either into the category “SMD” or the category “without SMD”. Therefore, there were no other exclusion criteria other than age < 20 years.

### Variable definitions

#### Outcomes

In Sweden, the first confirmed case of COVID-19 infection was reported on 31^st^ January 2020.^10^ The first COVID-19 related death was reported on 11^th^ March 2020.^11^ Our outcome was COVID-19 associated death, registered as such by the Swedish Board for Health and Welfare. We included all COVID-19 associated deaths occurring over a three-month period, from 11 March 2020 until 23 June 2020.

We analysed COVID-19 associated deaths as a dichotomous yes/no variable. The Swedish Board for Health and Welfare bases the criteria COVID-19 associated death on the underlying cause of death recorded on the death certificates. Two codes of the 10^th^ version of the International Classification of Diseases (ICD-10)^12^ are currently used, U07.1 or U07.2. Both codes fall into the ICD-10 chapter for provisional assignment of new diseases of uncertain aetiology or emergency use (U00 – U49). U07.1 is used when COVID-19 has been confirmed by laboratory testing irrespective of severity of clinical signs or symptoms. U07.2 is used when COVID-19 is diagnosed clinically or epidemiologically, but laboratory testing is inconclusive or not available.^12^

#### Exposures

The main exposure was SMD. We included bipolar or psychotic disorders with ICD-10 codes F20, F22, F25, F30 or F31 into the SMD variable. Individuals were included in the SMD category when there were at least two registered diagnoses between 1998 and 2019.

We also explored four somatic conditions available in the registers, diabetes, cardiovascular diseases, hypertension, and chronic respiratory diseases. We chose these conditions because they were thought to be relevant as risk factors for an adverse outcome at the beginning of the COVID-19 pandemic. We introduced a fifth category “none of the above”, which included all other individuals. We checked these somatic conditions for a time period from 2015-2019, based on diagnosis or pharmacological treatment received, using ICD and Anatomical Therapeutic Chemical Classification System (ATC) codes. We used the following ICD and ATC codes, (a) diabetes ICD E10-14, ATC A10, (b) cardiovascular diseases ICD I20-25, I48, I50, I61, I63, I64.9, I69.1, I69.3, I69.4, I69.8, I70, (c) hypertension ICD I10.9, I11-13, I15, ATC C02, C03, C07AB02, C08CA, C09, and (d) chronic respiratory diseases ICD J40-47, J60-67, J68.4, J70.1, J70.3, J96.1, J96.8, E 84.0. We stratified age into the following groups, 20-39, 40-59, 60-69, 70-79 and 80+ years.

### Statistical methods

The data was provided as summary data in anonymised form by the Swedish Board of Health and Welfare. Statistical analysis underlying this summary data was performed by a statistician at the National Board of Health and Welfare. The research group then analysed this data further. The data included information stratified according to case and controls on number of individuals in each age group and number of individuals in each risk factor category. From this data, we calculated the odds ratios (OR) for COVID-19 associated death for (a) the whole sample, (b) for each age group, and (c) for each risk factor category. For OR according to age-group, each death was only counted once. For OR according to risk factor category, a death could be counted several times when it appeared in more than one risk factor category. We calculated confidence intervals (CI) using Woolf’s formula for the standard error. Significance was tested with z-test. The significance level was set to 0.05 throughout, corresponding to a 95% CI. Where available, we compared the results of our z-test to the results of the Fisher exact test, provided by the National Board of Health and Welfare. Identification of significant relationships was 100% concordant.

#### Missing data

The number of deaths was available for all age groups. However, for some risk factor categories, the number of deaths had been withheld due to confidentiality reasons. For summary estimates of risk factors in the whole age group, we set missing data to 0.

### Ethics and consent procedures

The study was approved by the Swedish Ethical Review Authority (DNR 2020-02759) and conducted according to the declaration of Helsinki. The data originated from routine information collected by the Swedish Board of Health and Welfare, then made available as summary data in anonymised form for this first exploratory analysis. As only anonymised data was provided and potentially identifiable data was withheld, informed consent was not required.

## Results

### Baseline characteristics of the samples

The sample comprised of 7,923,859 individuals, 103,999 (1.3%) with SMD and 7,819,860 (98.7%) controls. There were significantly more individuals with SMD in the age groups between 40 and 69 years. There were significantly fewer individuals with SMD aged ≧ 70 years. Two risk factors under study, diabetes and chronic lung disease were significantly more common in individuals with SMD. Hypertension was marginally, but still significantly, more common in individuals with SMD. Cardiovascular disease was marginally, but still significantly, more common in the reference population (Table 1).

**Table 1:** Age and risk factor distribution in patients with severe mental disorders vs reference population.

|  | Population with severe<br>mental disorder<sup>a</sup><br>n = 103999 |  | Reference population<br>n = 7819860 |  |  |
| --- | --- | --- | --- | --- | --- |
|  | n | % | n | % | p |
| **Age groups** |  |  |  |  |  |
| 20-39 | 31246 | 30.0 | 2662638 | 34.0 | < 0.001 |
| 40-59 | 40899 | 39.3 | 2555319 | 32.7 | < 0.001 |
| 60-69 | 17163 | 16.5 | 1091275 | 14.0 | < 0.001 |
| 70-79 | 10986 | 10.6 | 978027 | 12.5 | < 0.001 |
| 80+ | 3705 | 3.6 | 532601 | 6.8 | < 0.001 |
| **Risk factors across all age groups<sup>bc</sup>** |  |  |  |  |  |
| Diabetes | 8012 | 7.7 | 327738 | 4.2 | < 0.001 |
| Cardiovascular<br>disease | 7308 | 7.0 | 573187 | 7.3 | < 0.001 |
| Hypertension | 10993 | 10.6 | 779557 | 10.0 | < 0.001 |
| Chronic lung<br>disease | 5664 | 5.4 | 231686 | 3.0 | < 0.001 |
| None of above | 83044 | 79.9 | 6599259 | 84.4 | < 0.001 |

n: number; p: p-value
<sup>a</sup>Diagnoses of severe mental disorders (bipolar or psychotic disorder) recorded between 1998 and 2019,<sup>b</sup>Diabetes, cardiovascular disease, hypertension, chronic lung disease, recorded between 2015 and 2019<sup>c</sup>Individuals could have more than one risk factor. Hence, the number of risk factors exceeded the number of individuals

### COVID-19 associated death

There were 130 (0.13%) deaths associated with COVID-19 infection in the SMD group and 4,945 (0.06%) in the control group. In the SMD group, 90.0% of COVID-19 diagnoses were ascertained by test (U07.1). In the reference group, 91.2 % of COVID-19 diagnoses were ascertained by test (U07.1). There were no significant differences in the proportion of COVID-19 diagnoses ascertained by test (U07.1) in both groups (p =0.419). The SMD group had double odds of COVID-19 associated death (OR 1.98, CI 1.66-2.35; p < 0.001). The odds were fourfold for death from COVID-19 infection for individuals in the age groups of 60-69 years (OR 4.52, CI 2.90-7.03; p < 0.001) and 70-79 years (OR 4.16, CI 3.14-5.52; p < 0.001). Regarding risk categories, higher OR were only found for cardiovascular diseases (OR 1.49, CI 1.11-1.99; p < 0.007). Individuals with SMD who had none of the sampled risk factors had threefold odds of COVID-19 associated death (OR 2.87, CI 2.15-3.83; <0,001) (Table 2).

**Table 2:** COVID-19 associated death having occurred between 1 January 2020 and 23 June 2020 in patients with severe mental disorder vs. reference population according to age *or* risk factors.

| Risk group | Population with<br>severe mental<br>disorder<br>n = 103999<sup>a</sup> |  | Reference<br>population<br>n = 7819860 |  |  |  |  |  |
| --- | --- | --- | --- | --- | --- | --- | --- | --- |
|  | n | % | n | % | OR | LCI | UCI | p |
| *Total sample* |  |  |  |  |  |  |  |  |
| N deaths | 130 | 0.13 | 4945 | 0.06 | 1.98 | 1.66 | 2.35 | <0.001 |
| *According to age group only* |  |  |  |  |  |  |  |  |
| 20-39 | - | - | - | - | _<sup>b</sup> | - | - | - |
| 40-59 | 6 | 0.01 | 167 | 0.01 | 2.24 | 0.99 | 5.07 | 0.052 |
| 60-69 | 21 | 0.12 | 296 | 0.03 | 4.52 | 2.90 | 7.03 | <0.001 |
| 70-79 | 51 | 0.46 | 1094 | 0.11 | 4.16 | 3.14 | 5.52 | <0.001 |
| 80+ | 52 | 1.40 | 3388 | 0.64 | 2.22 | 1.69 | 2.93 | <0.001 |
| *According to risk factor only<sup>c</sup>* |  |  |  |  |  |  |  |  |
| Diabetes | 32 | 0.40 | 1161 | 0.35 | 1.13 | 0.79 | 1.60 | 0.502 |
| Cardiovascular<br>disease | 46 | 0.63 | 2431 | 0.42 | 1.49 | 1.11 | 1.99 | 0.008 |
| Hypertension | 43 | 0.39 | 2644 | 0.34 | 1.15 | 0.85 | 1.56 | 0.353 |
| Chronic lung<br>disease | 13 | 0.23 | 633 | 0.27 | 0.84 | 0.48 | 1.46 | 0.533 |
| None of above | 48 | 0.06 | 1328 | 0.02 | 2.87 | 2.15 | 3.83 | <0.001 |

n: number; OR: odds ratio; LCI: lower 95% confidence interval; UCI: upper 95% confidence interval
<sup>a</sup>Diagnoses of severe mental disorders (bipolar or psychotic disorder) recorded between 1998 and 2018, risk factors recorded between 2015 and 2019
<sup>b</sup>OR not calculated because data withheld due to confidentiality reasons
<sup>c</sup>n Deaths counted for each risk factor. i.e. deaths associated with more than one risk factor will appear several times

## Discussion

### Main findings

To our knowledge, this is the first population-based study reporting on the risk of COVID- 19 associated mortality in individuals with SMD during the early months of the coronavirus outbreak. We found that individuals with SMD had almost double odds of COVID- 19 associated death compared to the reference population without SMD. Individuals with SMD aged between 60 and 79 years were particularly vulnerable with more than fourfold odds of COVID-19 associated death. Of the four risk factors available for study, cardiovascular disease increased the odds of COVID-19 associated death by 50%.

Individuals without any of the four risk factors under study, had threefold odds of COVID- 19 associated death.

### Severe mental disorder and death from other respiratory infections

At the time of writing, evidence regarding the association between SMD and COVID-19 infection is only emerging. Therefore, we cannot compare our findings directly with those of other studies. But our findings are in line with studies exploring the association between SMD and other (non-COVID-19) respiratory infections. A Swedish register study showed a three-to fourfold increased risk of death due to influenza or pneumonia in individuals with bipolar disorder.^1^ In individuals with schizophrenia, the risk was increased sevenfold.^2^ An American study also found a sevenfold increased risk of death due to pneumonia or influenza in adults with schizophrenia.^13^ A Danish register study explored all individuals hospitalised for any infection between 1995 and 2011. Individuals with bipolar or psychotic disorder had 52% increased mortality risk within 30 days after their infection.^14^ We could not find any study that explicitly explored risk factors associated with death from respiratory infections in individuals with SMD.

### Risk factors for a severe outcome from COVID-19 infection in the general population

The evidence regarding risk factors for a severe outcome from COVID-19 infection is rapidly evolving. The US Centers for Disease Control and Prevention (CDC) have collated a list of risk factors associated with severe illness from COVID-19 infection, defined as hospitalization, admission to the ICU, intubation or mechanical ventilation, or death. On this list, risk factors are rated into three categories according to quality of evidence, (a) strongest and most consistent evidence, (b) mixed evidence, and (c) limited evidence. Risk factors with the strongest and most consistent evidence include serious heart conditions, such as heart failure, coronary artery disease, or cardiomyopathies, cancer, chronic kidney disease, chronic obstructive pulmonary disease (COPD), obesity with a body mass index (BMI) ≥ 30, sickle cell disease, solid organ transplantation and type-2 diabetes mellitus. Risk factors with mixed evidence include asthma, cerebrovascular disease, hypertension, pregnancy, smoking, use of corticosteroids and other immunosuppressive medications. Risk factors with limited evidence include bone-marrow transplantation, HIV, immune deficiencies, inherited metabolic disorders, liver disease, neurological conditions, other chronic lung diseases, complex paediatric conditions, thalassaemia and type-1 diabetes mellitus.^15^ Notably, SMD is not mentioned in this list.

The CDC list identifies a large number of potential risk factors and comorbidities. Most of these could increase mortality risk in their own right. Thus, COVID-19 associated death may not necessarily be caused by COVID-19 infection. CDC statistics show that in just 6% of deaths involving COVID-19 infection, COVID-19 was the only cause mentioned. For the 94% deaths with conditions or causes in addition to COVID-19, there were on average 2.6 additional conditions or causes per death.^16^ An audit of 122 Covid-19 associated deaths from Östergötland County, Sweden, came to similar results. In only 15% of deaths, COVID- 19 infection was given as the direct cause. In 70% COVID-19 infection was thought to be a contributory factor but not the main cause. In the remaining 15%, the death could not be related to COVID-19 infection.^17^

### Risk factors for death from COVID-19 infection in individuals with severe mental disorder

In bipolar disorder and schizophrenia, comorbidity with at least one somatic condition is very common.^18,19^ When acute physically ill, individuals with SMD may then be sicker. One insurance claims study from Taiwan compared the risk of death in an intensive care unit (ICU) between 203 patients with schizophrenia and 2036 matched controls. In ICU, patients with schizophrenia were sicker, had a higher risk of acute organ dysfunction and death.^20^ Therefore, it is plausible that individuals with SMD may have a higher risk of COVID-19 associated death.

For our study, we chose four risk factors thought to be more prevalent in individuals with SMD.^13^ We chose these risk factors during the set-up of the study. At the time, evidence regarding risk factors was only emerging. Therefore, we made an informed guess that these four risk factors could affect the risk of COVID-19 associated mortality. But only cardiovascular disease led to a significantly increased OR in individuals with SMD. Cardiovascular conditions belong to the CDC category of risk factors with the strongest and most consistent evidence.^15^ For hypertension and chronic lung disease, we did not find an increased risk of death. Hypertension belongs to the CDC category of risk factors with mixed evidence.^15^ Chronic lung disease includes conditions that fall in the CDC categories of either mixed or limited evidence.^15^ Surprisingly, diabetes did not significantly increase the risk of COVID-19 associated death in our study. However, our study is only exploratory, sampling the first three months of the outbreak. There is also a chance that diabetes was underestimated in individuals with SMD. Despite rising awareness, diabetes may be one of the comorbidities easily missed in individuals with SMD.^21,22^

Our current analysis is exploratory, covering the first three months of the COVID-19 outbreak in Sweden. Therefore, the risk factor profile may change in future analyses. In our study, the OR was highest for the individuals with SMD who did not have any of our chosen four risk factors. This could suggest that SMD was an independent risk factor. However, there is no plausible mechanism which could explain a direct role of SMD in the pathophysiology of COVID-19 infection. More likely, there were other risk factors, not captured by our study, that increased the mortality risk from COVID-19 infection in individuals with SMD. Yet, this clear excess mortality risk makes individuals with SMD a risk group of their own, even if SMD *per se* is not involved in the pathophysiology of COVID-19 infections or its clinical course. Individuals with SMD should therefore be added to the groups already known to be at risk of serious illness from COVID-19 infection, i.e. the elderly, the obese or those with somatic comorbidities.

### Other risk factors which may specifically associated with severe mental disorder and/or its treatment

There may be other factors in individuals with SMD that can increase the risk for COVID-19 associated mortality. Such risk factors may either be associated with SMD itself, its pharmacological treatment or with a combination of both underlying SMD and its pharmacological treatment.

Risk factors associated with SMD include smoking and substance use disorder (SUD). Both remain highly prevalent in individuals with SMD.^23-25^ They increase the risk of pneumonia, cardiovascular disease, and compromised immunity. Increased risk for infection and subsequent worse outcomes may also result from difficulties to adhere to preventive measures.^26^ Such difficulties may become more prominent during periods of stress and isolation, which may precipitate relapse into an acute SMD episode.^27,28^

Risk factors associated with the pharmacological treatment include medication associated pneumonia, neutropenia and QT prolongation. Exposure to first-generation antipsychotics (FGA) or second-generation antipsychotics (SGA) may double the risk of pneumonia.^29^ Mortality from pneumonia may also increase.^30^ Mood stabilisers such as valproate and carbamazepine may be risk neutral. Lithium may be protective,^31,32^ for reasons yet to be explained. Benzodiazepines and benzodiazepine related drugs (BZRD), taken by 30% to 60% of individuals with schizophrenia or bipolar disorder, are other concerns.^33-36^ Neutropenia and its extreme form agranulocytosis can occur with a variety of antipsychotics and mood-stabilisers, particularly with clozapine and carbamazepine.^37^ Some of the agents used to treat COVID-19 infection such as chloroquine, hydroxychloroquine, azithromycin, and lopinavir can also cause neutropenia.^38^ Hence, individuals with SMD taking such agents need careful monitoring.^39^ QT prolongation is a potentially dangerous adverse effect increasing the risk of torsade de pointes and current cardiac death. Many antipsychotics can prolong QT interval. Citalopram, escitalopram, tricyclic antidepressants and methadone can also prolong the QT interval. Intravenous administration, combination therapy or excess doses also increase the risk for QT prolongation.^37^ These psychotropic agents may become problematic in combination with somatic drugs also increasing QT interval, used for treating a COVID-19 infection. The latter group includes some antibiotics and antiarrhythmics, chloroquine, hydroxychloroquine and the antiviral and histamin-2 antagonist famotidine.^38^ It currently remains unclear how often such interactions with psychotropic drugs occur in the context of COVID-19 treatment.

Risk factors associated with the underlying SMD and its treatment include obesity and venous thromboembolism (VTE). The likelihood of obesity is 2.8 to 4.4 times increased in individuals with schizophrenia and about 1.2 to 1.7 times increased in individuals with bipolar disorder or major depression.^40^ In part, this increased risk is associated with psychotropic medications. Antipsychotics are of particular concern. Of all antipsychotics, clozapine and olanzapine have the highest risk of weight gain.^37,40^ Both schizophrenia and bipolar disorder are associated with an increased risk venous thromboembolism (VTE) in form of deep-vein thrombosis (DVT) or pulmonary embolism (PE). This increased risk of venous thromboembolism may be related to a higher risk of smoking and obesity in individuals with SMD. Immobilisation, including lack of exercises, restraints and lower leg paralysis, and treatments with antipsychotics may constitute further risk factors for VTE.^41,42^ Antipsychotics have also been implicated to increase the risk of VTE. Risk estimates range from 50% to threefold increased risk, depending on substance class.^43,44^

### Strengths

The major strength of this study is its large sample-size in a naturalistic setting. The study is representative with register data covering the entire Swedish population aged 20 years and older. Therefore, there is no scope for selection bias. Individuals fell into one group (SMD) or the other group (reference population); no further exclusion criteria were warranted. The summary data was prepared independently from the research group by a statistician at the Swedish Board of Health and Welfare. Hence, the scope for observation bias was minimised. A further strength lies in the accuracy of Swedish register data. For instance, the Swedish Cause of Death Register covers more than 99% of all deaths.

### Limitations

Our data covered the early months of the COVID-19 outbreak in Sweden. In this emerging situation the absolute number of deaths was still relatively small, which could affect power. In order to maximise power, we amalgamated bipolar and psychotic disorders into SMD as one exposure category. Several other examples of epidemiological studies exist, where mood and psychotic disorders are amalgamated in similar ways.^3,14,41,45-47^ Here, we combined psychotic and bipolar disorders into one category because the prevalence for somatic comorbidities and excess mortality are similar for both conditions.^1,2^ We did not include severe depression in our SMD variable, since severe depression is a more heterogenous group. For this group, based on register data alone, it can be difficult to establish whether depression is the cause or consequence of somatic comorbidity. Excluding individuals with severe depression may most likely have led to underestimate but not an overestimate of the risk of COVID-19 associated mortality for individuals with SMD.

For this exploratory analysis, we had to rely on summary data. Summary data are much less detailed than individual level data. However, we decided to report our summary data at this point to alert clinicians to this new risk group. We intend to conduct further analyses with individual level data as soon as possible. With individual level data it will be possible to adjust for baseline variables, such as age and sex, psychotropic drugs used and other variables of potential importance as outlined in our discussion. The association between COVID-19 associated mortality and SMD involves most likely a large quantity of biological and psychosocial factors. Some of these will be confounders, but others may actually lie on exposure-outcome causal pathways. Thus, even though our finding that SMD doubles the odds of COVID-19 associated death is crude and preliminary, it is nevertheless noteworthy. To further study the impact of SMD on the risk of COVID-19 infection or associated deaths, both register and clinical studies are needed. Register studies have the advantage of large sample sizes, but clinical information is limited. Clinical studies will be smaller, but risk factors can be explored in more detail.

In the beginning of the pandemic, testing was not ubiquitously available. Thus, diagnoses were also made clinically. This is reflected by the provisional ICD code U07.2. Thus, there was scope for misclassification due to false positive COVID-19 diagnoses. In both groups, about 90% of diagnoses were confirmed by laboratory testing (U07.1). Hence, the scope for misclassification is comparably low in both groups. As acknowledged earlier in this discussion, not every death associated with COVID-19 infection may have been caused by COVID-19 infection.^16,17^ Distinguishing between causation and association may have been particularly difficult in the early months of the COVID-19 outbreak. However, this would have affected cases and controls in the same way without any impact on the OR.

### Conclusions

Our preliminary results suggest that individuals with SMD may be a further group at increased risk of COVID-19 associated death. It is important that clinicians are alerted to this new risk group. This increased mortality can be associated with a higher prevalence of somatic morbidity and life-style related risk factors. These factors require clarification to enable clinicians to provide the best physical and mental health care tailored to special requirements of this risk group.

## Data Availability

The summary tables provided by the Swedish Board of Health and Welfare are already included in this article. Requests for the original summary tables provided will be taken up with Swedish Board of Health and Welfare.

## Declarations

## Acknowledgement

We gratefully acknowledge Olga Filson for her assistance with the data analysis.

## Ethics statement

The study was approved by the Swedish Ethical Review Authority (DNR 2020-02759) and conducted according to the declaration of Helsinki. The data originated from routine information collected by the Swedish Board of Health and Welfare, which was then made available as summary data in anonymised form. As this data was only provided in anonymised form, informed consent was not required. Potentially identifiable data was withheld by the Swedish Board of Health and Welfare (data withheld due to confidentiality reasons).

## Author contribution

MM, UW, MB, LÖ, MW: conception and design of the work. MM, UW; LÖ: acquisition and analysis of data. UW, MM, MB: drafting the work. MM, UW, MB, LÖ, MW: revising and providing the final approval of the work.

